# A novel cortical biomarker signature for predicting pain sensitivity: protocol for the PREDICT longitudinal analytical validation study

**DOI:** 10.1101/2020.02.18.20024695

**Authors:** David A Seminowicz, Katarzyna Bilska, Nahian S Chowdhury, Patrick Skippen, Samantha K Millard, Alan KI Chiang, Shuo Chen, Andrew J Furman, Siobhan M Schabrun

## Abstract

**Introduction:** Temporomandibular disorder (TMD) is a common musculoskeletal pain condition with development of chronic symptoms in 49% of patients. Although a number of biological factors have shown an association with chronic TMD in cross-sectional and case control studies, there are currently no biomarkers that can predict the development of chronic symptoms. The PREDICT study aims to undertake analytical validation of a novel peak alpha frequency (PAF) and corticomotor excitability (CME) biomarker signature using a human model of the transition to sustained myofascial temporomandibular pain (masseter intramuscular injection of nerve growth factor [NGF]). This paper describes, *a-priori*, the methods and analysis plan.

**Methods and analysis:** This study uses a multi-site longitudinal, experimental study to follow individuals for a period of 30 days as they progressively develop and experience complete resolution of NGF-induced muscle pain. 150 healthy participants will be recruited. Participants will complete twice daily electronic pain dairies from Day 0 to Day 30 and undergo assessment of pressure pain thresholds, and recording of PAF and CME on Days 0, 2 and 5. Intramuscular injection of NGF will be given into the right masseter muscle on Days 0 and 2. The primary outcome is pain sensitivity.

**Ethics and dissemination:** Ethical approval has been obtained from The University of New South Wales (HC190206) and the University of Maryland Baltimore (HP-00085371). Dissemination will occur through presentations at National and International conferences and publications in international peer-reviewed journals.

**Registration details:** ClinicalTrials.gov: NCT04241562 (prospective)

**STRENGTHS AND LIMITATIONS OF THIS STUDY:**

- PREDICT is the first study to undertake analytical validation of a peak alpha frequency and corticomotor excitability biomarker signature. The study will determine the sensitivity, specificity and accuracy of this biomarker signature at predicting pain sensitivity.
- PREDICT will establish the reportable range of test results and determine automation and simplification of methods for biomarker detection in the clinic.
- The methods and statistical analysis plan are pre-specified to ensure reporting transparency.
- Future patient studies will be required for clinical validation.

## INTRODUCTION

Temporomandibular disorder (TMD) is the second most common musculoskeletal pain condition after back pain, with an annual incidence of 4% and development of chronic symptoms in 49% of patients^34, 36^. Although a number of biological factors have shown an association with chronic TMD in cross-sectional and case control studies including sensitivity to mechanical stimuli^37^, up-regulated central nociceptive processing^29, 31^, increased heart rate and reduced heart rate variability^22^, single nucleotide polymorphisms^35, 38^, elevated levels of pro-inflammatory cytokines^35^, elevated interstitial glutamate concentration^2^, and altered brain structure and function^21^, these have either failed to yield clinically meaningful predictive power or have not undergone comprehensive validation in prospective trials. Consequently, there are no biomarkers available that can predict the development of chronic TMD. In fact, there are no biomarkers qualified (considered valid and psychometrically sound) by the FDA for use in clinical trials or clinical practice for any musculoskeletal pain condition^39^.

In most patients with chronic musculoskeletal pain a peripheral anatomical cause for pain cannot be identified. For example, myofascial TMD is more commonly associated with stress and anxiety than anatomical pathology^40^ while 90% of all chronic low back pain is diagnosed as ‘non-specific’^18^. In conditions where a structural impairment can be detected (i.e. articular cartilage damage in osteoarthritis), the magnitude of pain fails to correlate with the extent of tissue damage^11^. These observations suggest a role for the brain in the development and maintenance of chronic pain. Indeed, early investigations suggest that variability in brain connectivity circuits can predict sensitivity to a transient pain stimulus in healthy individuals^30^. Although these data have not yet been expanded and the relevance to clinical pain is unknown, brain imaging methods are widely considered to have potential as diagnostic, prognostic and predictive biomarkers of chronic pain^5^.

Using brain imaging methods of electroencephalography (EEG) and transcranial magnetic stimulation (TMS), preliminary evidence for a unique biomarker signature – combined resting state peak alpha frequency (PAF) and corticomotor excitability (CME) – has recently been demonstrated. In studies using long-lasting human pain models, slow PAF and low CME are associated with high pain severity and longer pain duration^13, 32, 33^. Consistent with this, low CME in the acute stage of clinical pain is associated with high pain severity and the presence of pain at 6 months follow-up^3^. These data suggest the combination of slow PAF and low CME may be a plausible predictive biomarker for the development of chronic TMD.

Here, we outline the experimental protocol and statistical analysis plan to undertake extensive analytical validation of the PAF/CME biomarker signature using a standardized human model of the transition to sustained myofascial temporomandibular pain (masseter intramuscular injection of nerve growth factor). We hypothesise that the PAF/CME biomarker signature will predict pain sensitivity (primary) and pain severity and duration (secondary) with at least 75% accuracy in a human transitional pain model of TMD. In addition, we aim to i) determine the sensitivity, specificity and accuracy of the PAF/CME biomarker at predicting pain sensitivity, severity, and duration, ii) determine the reportable range of test results and reference intervals for fast vs. slow PAF and high vs. low CME and iii) establish optimization of the model and automation and simplification of methods for biomarker detection.

## METHODS

### Design

A multi-site longitudinal, experimental study will be used to follow healthy individuals for a period of 30 days as they progressively develop and experience complete resolution of NGF-induced muscle pain. All data collection will be performed at the Australian site (Neuroscience Research Australia; NeuRA), and blinded data processing and analyses will be performed at the USA site (the University of Maryland Baltimore; UMB). The UMB site will also be responsible for standardization and automation of analytical methods. A data and safety monitoring plan has been established and an independent monitoring committee will conduct annual reviews of study progress and safety. Ethical approval has been obtained from the University of New South Wales (HC190206) and the University of Maryland Baltimore (HP-00085371). All procedures will be conducted in accordance with the Declaration of Helsinki. Written, informed consent will be obtained and participants will be free to withdraw from the study at any time. The study is prospectively registered on ClinicalTrials.gov (NCT04241562).

### Patient and public involvement

Patients and the public were not involved in the design of this protocol. Individual results will be provided to participants on request and a summary of the overall outcomes of the study will be available to all participants on completion of the trial.

### Participants

### Inclusion and exclusion criteria

Healthy men and women with no medical complaints, no history of chronic pain and no current acute pain between the ages of 18 and 44 years will be included. These inclusion criteria are justified based on data from the OPPERA prospective cohort study that demonstrates only marginally greater TMD incidence in females than males and an incidence rate of first-onset TMD of 2.5% per annum among 18 to 24-year olds and 4.5% per annum among 35 to 44-year olds^34^. Exclusion criteria are: 1) inability or refusal to provide written consent, 2) presence of any acute pain disorder, 3) history or presence of any chronic pain disorder, 4) history or presence of any other medical or psychiatric compliant, 5) use of opioids or illicit drugs in the past 3 months, 6) pregnant or lactating women, 7) contraindicated for TMS (metal implants, epilepsy)^16^. Participants will be recruited via notices placed on community notice boards at UNSW and NeuRA, flyers, mailings and social media platforms (such as Facebook) as well as the use of a volunteer healthy participant database held by NeuRA.

### Sample size

150 healthy subjects will be included. Our preliminary data indicate consistent associations between PAF and future pain severity, as well as strong relationships between CME and pain severity. The design of the current discovery-based study is not amenable to traditional power calculations because the outcomes are not p-value-based inference but rather predictive. Larger sample sizes in the training set give better classification, while larger sample sizes in the testing set give higher accuracy. We have chosen a sample size that provides good classification and accuracy. Allowing for a 10% drop-out rate, we will enrol 165 subjects.

### Data collection procedures

### Overview

Participants will first complete a phone screen and if eligible, a time will be made for the Day 0 visit. At the Day 0 visit, after reviewing eligibility criteria, participants will complete informed consent (considered enrolment in the study) and questionnaires. Participants will complete twice daily electronic pain dairies from Day 0 to Day 30 and attend 3 laboratory visits of ∼ 2 hours duration on Days 0, 2 and 5. Each laboratory visit will include assessment of pressure pain thresholds (PPTs), and recording of PAF and CME. Intramuscular injection of NGF will be given into the right masseter muscle at the end of each test session on Days 0 and 2 (Fig 1). These procedures are detailed below.

**Figure 1.**
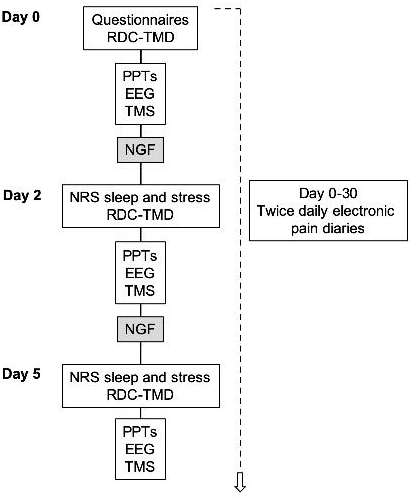
Experimental protocol. Questionnaires – Health history form, Pain Catastrophising Scale, Brief Pain Inventory Pain Severity and 7-item Interference subscales; SF-8 Health Survey; Sleep Scale; Patient Health Questionnaire-2 item; Generalised Anxiety Disorder 2 item Questionnaire; Tobacco, Alcohol, Prescription Medications, and other Substances Questionnaire, Perceived Stress Scale. Abbreviations: RDC-TMD - Research Diagnostic Criteria questionnaire for temporomandibular disorder, PPTs – Pressure Pain Thresholds, EEG – Electroencephalography, TMS – Transcranial Magnetic Stimulation, NGF – Nerve growth factor, NRS – Numerical rating scale.

### Electronic diaries

Diaries will be completed using a computer, tablet or phone at 10am and 7pm each day from Day 0 to Day 30. Electronic diary completion will take 2-mins. Participants will rate their pain intensity on an 11-point numerical rating scale (NRS) anchored with ‘no pain’ at zero and ‘worst pain imaginable’ at 10 at rest, and while chewing, swallowing, drinking, talking, yawning and smiling^8^. Study staff will send a text message containing a unique link to the pain dairy synced with the time of diary completion (10am and 7pm) each day. If the diary is not completed for two consecutive days, participants will be followed-up by phone.

### Questionnaires

At Day 0 only, participants complete a health history form assessing medical history. We will use the following National Institutes of Health common data elements (CDE) for pain biomarkers including: Pain Catastrophizing Scale (PCS)^41^; Brief Pain Inventory (BPI) Pain Severity and 7-item Interference subscales^17, 43^; SF-8 to assess general health^48^; Sleep Scale; PHQ-2 to assess depression^19^; GAD-2 to assess anxiety^20^; Tobacco, Alcohol, Prescription medications, and other Substances (TAPS). Participants will also complete the Perceived Stress Scale (PSS)^44^. These questionnaires assess factors that have been associated with first onset and/or chronic TMD^1^ and often worsen as TMD progresses^10^. These questionnaires will be completed on Day 0.

On Days 0, 2 and 5 participants will complete the Research Diagnostic Criteria questionnaire for TMD^9^ and two NRS scales asking the following questions:

On a scale of 1 to 10 where 1 is ‘poor sleep quality’ and 10 is ‘excellent sleep quality’, how would you rate your sleep last night?

On a scale of 1 to 10 where 1 is ‘not at all stressed’ and 10 is ‘very stressed’, how would you rate your level of stress over the last 24 hours?

### Pressure pain Thresholds (PPTs)

Because NGF injection is known to sensitize mechanosensitive afferents^23, 42^, and because lower PPTs at cranial sites are associated with increased risk of developing TMD^14^ and fluctuate with the clinical disease course^37^, we will assess PPTs at 5 sites - overlying the masseter muscle, temporalis muscle, the temporomandibular joint, the trapezius muscle, and the lateral epicondyle. Three measures will be made at each site, with 1 min rest between measurements at the same site, in pseudorandomized order using a commercially available algometer.

### Peak alpha frequency (PAF)

Scalp EEG will be collected using Brain Vision actiCAP with at least 32-channels, following the extended international 10–20 system^28^, a BrainAmp DC amplifier and Brain Vision Recorder version 1.22.0101 software (all Brain Products GmbH, Munich, Germany). Auxiliary recordings will include skin conductance, respiration, and electrocardiogram (ECG). Participants will be asked to make facial muscle contractions such as clenching their teeth, blinking, and saccades, while EEG is recorded. This will take about two minutes and will be used to aid in automated artefact removal. Participants will then be told to relax their muscles and resting state eyes closed EEG will be recorded for 5 minutes and used for PAF calculation.

#### Corticomotor excitability

Rapid TMS will be used to map the primary motor cortical representation of the right masseter muscle and right extensor carpi radialis brevis (ECRB) muscle. Mapping of the right ECRB muscle is included to determine whether any changes in corticomotor excitability are restricted to the affected muscle. Single-pulse, biphasic stimuli will be delivered to the left hemisphere using a Magstim Super Rapid^2^ Plus and a 70mm figure of eight coil. Bipolar surface electrodes will be used to record electromyographic (EMG) activity^7^. EMG signals will be amplified (x2000), filtered (20-1000 Hz) and digitally sampled at 5kHz. The scalp site that evokes the largest EMG response (motor evoked potential, MEP) at a given TMS intensity will be determined for each muscle in each individual (termed the ‘hotspot’) and the active (aMT – masseter muscle) or resting (rMT – ECRB muscle) motor threshold calculated. A 6 × 6 cm grid will be generated in the neuronavigation software for each muscle, centred to each participant’s hotspot. 110 stimuli will be delivered at 2-sec intervals to pseudorandom locations over the grid at 120% of aMT for the masseter muscle and 120% of rMT for the ECRB muscle.

### Intramuscular injection of nerve growth factor (NGF)

After cleaning the skin with alcohol, a sterile solution of recombinant human NGF (dose of 5 μg [0.2 ml]) will be given as a bolus injection into the muscle belly of the right masseter on days 0 and 2 using a 1-ml syringe with a disposable needle. Any individual who does not develop sensitivity to the NGF model, assessed by diary pain ratings and pressure pain thresholds of the injected muscle, will be considered a non-responder and excluded from analyses.

## Outcome measures

### Primary Outcome

The primary outcome is pain sensitivity: participants are dichotomized as high- or low-pain sensitive based on the peak pain severity from diary recordings^12, 33^. That is, based on pain severity in the training set (n=100), participants will be classified as the top 40% high- or bottom 40% low-pain sensitive. This classification can be further weighted (e.g. very high, very low) as described in Aim 1.3.

### Secondary Outcomes

The secondary outcomes are pain severity (peak average daily pain severity based on diaries on a 0-10 scale) and pain duration, defined as the time between pain onset and complete resolution of pain (0 on a 0-10 scale for two consecutive days).

### Biomarker candidates

Biomarker candidates are PAF at Day 0 and CME at Day 5. As this is a discovery project, we will also examine PAF and CME at every day it is tested (see Aim 1.3 below).

## Data processing

### PAF and other EEG metrics

All data processing will be performed using custom MATLAB scripts implementing EEGLAB^6^ and FieldTrip toolboxes^27^. Data will be referenced to the average across all recording channels and segmented into 5-second epochs. These epochs are manually inspected and all epochs containing marked muscular artifacts are rejected. Channels with poor recordings will be rejected. Principal component analysis is then applied to identify and remove components relating to eye blinks, saccades, and ECG artifacts. Power spectral density will be derived in .20 Hz bins and the 2-40 Hz range will be extracted. Power spectral density will be extracted in sensor space around sensorimotor cortices (C3, Cz, C4, and neighboring electrodes), as well as sensorimotor ICA components demonstrating clear alpha peaks^26, 46, 47^. A Hanning taper will be applied to the data prior to calculating the spectra to reduce any edge artifacts similar to the approach taken in^24-26^. PAF is calculated using the center of gravity method, as we have done previously^12^.

### Corticomotor excitability

All data processing is performed using a custom MATLAB script. Triangular linear interpolation is used to create a full surface map within a transformed 2D plane containing the stimulation coordinates and their corresponding peak-to-peak MEP amplitudes^4, 45^. The resultant map is divided into 2500 partitions (50 × 50), with each partition assigned an approximated value based on the nearest acquired MEP data. Map area is determined as the ratio of the number of approximated partitions where the MEP exceeds 10% of the maximum MEP across all partitions. This cut-off reduces data variability. Map volume is then calculated as the sum of all MEPs (subtracted by the 10% level). This approach is described in full detail (including relevant equations) here^45^.

## Statistical analyses

### Aim 1.1: Predicting pain sensitivity and optimizing the model

We will validate the PAF/CME biomarker signature and test the predictive accuracy using a nested control-test scheme. The sample of 150 subjects will first be randomly divided into an outer-training set (n = 100) and an outer-testing set (n = 50). The research team at UMB will be blinded to the outcomes of pain sensitivity, severity and duration in the outer-testing cohort. “High pain sensitive” subjects are defined as the 40% of all subjects with the highest pain sensitivity, whereas “low pain sensitive” subjects are the 40% of all subjects with the lowest pain sensitivity. The ratios of high-vs. low-pain sensitive individuals will be matched between the two cohorts. Next, the outer-training cohort will be split into 5-folds (20 subjects for each fold) for cross validation. Each fold of 20 subjects will be tested as an inner testing cohort based on the remaining 4 folds as the inner training cohorts. We expect the 5-fold cross validation will provide sound performance assessment with balanced variance-bias trade-off (see details in^15^). We will consider multiple classifiers including logistic regression, support vector machine, gradient boosting, random forest, and neural networks. These predictive models along with the tuning parameters will be compared based on the performance of the 5-fold cross validation. The biomarkers may predict outcomes in a nonlinear fashion, and thus most machine learning models (e.g. support vector machine and gradient boosting+random forest) will detect nonlinear functions. The predictive model with the highest performance (i.e., the final model) based on the ability to classify the 40% most pain sensitive and the 40% least pain sensitive participants will be referred to as the “winning classifier”. The parameters of the winning classifier will be fixed and used to predict the outcomes of the outer-testing set. After finalizing the predicted outcomes, the outcomes will be unblinded to the UMB team. We will compare the predicted outcomes with the true outcomes and assess the accuracy, sensitivity, specificity, positive and negative predictive values. The predictive accuracy based on binary outcome prediction is used because it is more robust than mean squared error of a predictive model for continuous variables and is more commonly used in the field. Our target is to achieve an area under the curve of the receiver operating characteristic greater than or equal to 75% when applying the fixed classifier to the testing data set.

### Aim 1.2: Reportable ranges

The sensitivity, specificity and accuracy of the PAF/CME biomarker will be based on the blinded prediction of the outer-training 50 samples. Reference intervals will be reported for the whole sample, including intervals for fast vs. slow PAF and high vs. low CME. These will be reported as tables, standardized by age, sex, and other factors. We will further report on the stability of these measures over time (Days 0, 2 and 5).

### Aim 1.3: Optimization

We will explore how the inclusion of other combinations of factors in the model affects performance characteristics. The auxiliary factors considered in the model will include questionnaire and diary data, PPTs, and other EEG data (theta, alpha, beta, low gamma power) using a model/variable selection procedure to further boost the performance of the model. The nested training-testing scheme will be used to determine the optimal pain sensitivity prediction model using the biomarkers.

Weighted accuracy: since the low- and high-pain sensitive categories are determined based on a continuous pain scale, subjects with pain intensities near the median should be weighted less. Therefore, in addition to the simple accuracy, the weighted accuracy will be calculated. The weight will be determined by the distance of pain levels to the high-low cut-off.

### Automation and Simplification of Methods

In order for the biomarker signature to have application to large populations and settings, users must be able to rapidly collect and analyze data with minimal training. We will develop methods that automatically produce biomarker readouts with minimal human input, thus reducing bias associated with data input. Our goal will be to develop a method for automated signature calculation that achieves an intraclass coefficient of at least 80% compared to output from non-automated data processing and no significant difference between automated and non-automated based on bootstrap inference.

## Data Availability

De-identified, individual participant data will be made available immediately following publication via an open-access data repository.

## ETHICS AND DISSEMINATION

Ethical approval has been obtained from The University of New South Wales (HC190206) and the University of Maryland Baltimore (HP-00085371). Dissemination will occur through presentations at National and International conferences and publications in international peer-reviewed journals.

## Funding

This project is funded by grant 1R61NS113269-01 from The National Institutes of Health to DAS, SMS and SC. SMS receives salary support from The National Health and Medical Research Council of Australia (1105040). SKM and KB hold University of New South Wales International Postgraduate Scholarships.

### Author contributions

DAS, SMS and SC acquired funding to undertake this research. DAS, SMS and KB drafted the protocol. DAS, SMS, KB, AC, NC, AF, SKM, PS, and SC contributed to revisions and approved the final version of the manuscript.

## REFERENCES

1. Bair E, Gaynor S, Slade GD, Ohrbach R, Fillingim RB, Greenspan JD, Dubner R, Smith SB, Diatchenko L, Maixner W. Identification of clusters of individuals relevant to temporomandibular disorders and other chronic pain conditions: the OPPERA study. Pain. 157:1266-1278, 2016

2. Castrillon EE, Ernberg M, Cairns BE, Wang K, Sessle BJ, Arendt-Nielsen L, Svensson P. Interstitial glutamate concentration is elevated in the masseter muscle of myofascial temporomandibular disorder patients. J Orofac Pain. 24:350-360, 2010

3. Chang WJ, Buscemi V, Liston MB, McAuley JH, Hodges PW, Schabrun SM. Sensorimotor Cortical Activity in Acute Low Back Pain: A Cross-Sectional Study. J Pain. 2019

4. D’Errico J: Surface fitting using gridfit. MATLAB central file exchange. Available at: https://au.mathworks.com/matlabcentral/fileexchange/8998-surface-fitting-using-gridfit xAccessed 1 June, 2017

5. Davis KD, Flor H, Greely HT, Iannetti GD, Mackey S, Ploner M, Pustilnik A, Tracey I, Treede RD, Wager TD. Brain imaging tests for chronic pain: medical, legal and ethical issues and recommendations. Nat Rev Neurol. 13:624-638, 2017

6. Delorme A, Makeig S. EEGLAB: an open source toolbox for analysis of single-trial EEG dynamics including independent component analysis. J Neurosci Methods. 134:9-21, 2004

7. Deng H, Gao S, Lu S, Kumar A, Zhang Z, Svensson P. Alteration of occlusal vertical dimension induces signs of neuroplastic changes in corticomotor control of masseter muscles: Preliminary findings. J Oral Rehabil. 45:710-719, 2018

8. Dworkin SF. Research Diagnostic criteria for Temporomandibular Disorders: current status & future relevance. J Oral Rehabil. 37:734-743, 2010

9. Dworkin SF, LeResche L. Research diagnostic criteria for temporomandibular disorders: review, criteria, examinations and specifications, critique. J Craniomandib Disord. 6:301-355, 1992

10. Fillingim RB, Slade GD, Greenspan JD, Dubner R, Maixner W, Bair E, Ohrbach R. Long-term changes in biopsychosocial characteristics related to temporomandibular disorder: findings from the OPPERA study. Pain. 159:2403-2413, 2018

11. Finan PH, Buenaver LF, Bounds SC, Hussain S, Park RJ, Haque UJ, Campbell CM, Haythornthwaite JA, Edwards RR, Smith MT. Discordance between pain and radiographic severity in knee osteoarthritis: findings from quantitative sensory testing of central sensitization. Arthritis Rheum. 65:363-372, 2013

12. Furman AJ, Krimmel, J., Zhang, M., Keaser, R., Gullapalli, D., Seminowicz, D.A.,. The relationship of Sensorimotor Peak Alpha Frequency to regions across the brain is modulated by pain. No. 391.21. 2018 Neuroscience Meeting Planner. San Diego, CA: Society for Neuroscience, 2018. 2018

13. Furman AJ, Thapa T, Summers SJ, Cavaleri R, Fogarty JS, Steiner GZ, Schabrun SM, Seminowicz DA. Cerebral peak alpha frequency reflects average pain severity in a human model of sustained, musculoskeletal pain. J Neurophysiol. 122:1784-1793, 2019

14. Greenspan JD, Slade GD, Bair E, Dubner R, Fillingim RB, Ohrbach R, Knott C, Diatchenko L, Liu Q, Maixner W. Pain sensitivity and autonomic factors associated with development of TMD: the OPPERA prospective cohort study. J Pain. 14:T63-74 e61-66, 2013

15. Hastie T, Tibshirani, R., Friedman, J.,: The elements of statistical learning, second edition, Springer-Verlag, New York, 2009.

16. Keel JC, Smith MJ, Wassermann EM. A safety screening questionnaire for transcranial magnetic stimulation. Clin Neurophysiol. 112:720, 2001

17. Keller S, Bann CM, Dodd SL, Schein J, Mendoza TR, Cleeland CS. Validity of the brief pain inventory for use in documenting the outcomes of patients with noncancer pain. Clin J Pain. 20:309–318, 2004

18. Koes BW, van Tulder MW, Thomas S. Diagnosis and treatment of low back pain. BMJ. 332:1430–1434, 2006

19. Kroenke K, Spitzer RL, Williams JB. The Patient Health Questionnaire-2: validity of a two-item depression screener. Med Care. 41:1284-1292, 2003

20. Kroenke K, Spitzer RL, Williams JB, Monahan PO, Lowe B. Anxiety disorders in primary care: prevalence, impairment, comorbidity, and detection. Ann Intern Med. 146:317-325, 2007

21. Lin CS. Brain signature of chronic orofacial pain: a systematic review and meta-analysis on neuroimaging research of trigeminal neuropathic pain and temporomandibular joint disorders. PLoS One. 9:e94300. 2014

22. Maixner W, Greenspan JD, Dubner R, Bair E, Mulkey F, Miller V, Knott C, Slade GD, Ohrbach R, Diatchenko L, Fillingim RB. Potential autonomic risk factors for chronic TMD: descriptive data and empirically identified domains from the OPPERA case-control study. J Pain. 12:T75-91, 2011

23. Mann MK, Dong XD, Svensson P, Cairns BE. Influence of intramuscular nerve growth factor injection on the response properties of rat masseter muscle afferent fibers. J Orofac Pain. 20:325-336, 2006

24. Mazaheri A, Coffey-Corina S, Mangun GR, Bekker EM, Berry AS, Corbett BA. Functional disconnection of frontal cortex and visual cortex in attention-deficit/hyperactivity disorder. Biol Psychiatry. 67:617-623, 2010

25. Mazaheri A, Nieuwenhuis IL, van Dijk H, Jensen O. Prestimulus alpha and mu activity predicts failure to inhibit motor responses. Hum Brain Mapp. 30:1791-1800, 2009

26. Mazaheri A, van Schouwenburg MR, Dimitrijevic A, Denys D, Cools R, Jensen O. Region-specific modulations in oscillatory alpha activity serve to facilitate processing in the visual and auditory modalities. Neuroimage. 87:356-362, 2014

27. Oostenveld R, Fries P, Maris E, Schoffelen JM. FieldTrip: Open source software for advanced analysis of MEG, EEG, and invasive electrophysiological data. Comput Intell Neurosci. 2011:156869, 2011

28. Oostenveld R, Praamstra P. The five percent electrode system for high-resolution EEG and ERP measurements. Clin Neurophysiol. 112:713-719, 2001

29. Raphael KG, Janal MN, Anathan S, Cook DB, Staud R. Temporal summation of heat pain in temporomandibular disorder patients. J Orofac Pain. 23:54-64, 2009

30. Rogachov A, Cheng JC, Erpelding N, Hemington KS, Crawley AP, Davis KD. Regional brain signal variability: a novel indicator of pain sensitivity and coping. Pain. 157:2483-2492, 2016

31. Sarlani E, Grace EG, Reynolds MA, Greenspan JD. Evidence for up-regulated central nociceptive processing in patients with masticatory myofascial pain. J Orofac Pain. 18:41-55, 2004

32. Seminowicz DA, Thapa T, Schabrun SM. Corticomotor Depression is Associated With Higher Pain Severity in the Transition to Sustained Pain: A Longitudinal Exploratory Study of Individual Differences. J Pain. 2019

33. Seminowicz DA, Thapa, T., Furman, A., Summers, S., Cavaleri, R., Fogarty, J., Steiner, G., Schabrun, SM.,. Slow peak alpha frequency and corticomotor depression linked to high pain susceptibility in transition to sustained pain. bioRXiv. 2017

34. Slade GD, Bair E, Greenspan JD, Dubner R, Fillingim RB, Diatchenko L, Maixner W, Knott C, Ohrbach R. Signs and symptoms of first-onset TMD and sociodemographic predictors of its development: the OPPERA prospective cohort study. J Pain. 14:T20-32 e21-23, 2013

35. Slade GD, Conrad MS, Diatchenko L, Rashid NU, Zhong S, Smith S, Rhodes J, Medvedev A, Makarov S, Maixner W, Nackley AG. Cytokine biomarkers and chronic pain: association of genes, transcription, and circulating proteins with temporomandibular disorders and widespread palpation tenderness. Pain. 152:2802–2812, 2011

36. Slade GD, Ohrbach R, Greenspan JD, Fillingim RB, Bair E, Sanders AE, Dubner R, Diatchenko L, Meloto CB, Smith S, Maixner W. Painful Temporomandibular Disorder: Decade of Discovery from OPPERA Studies. J Dent Res. 95:1084–1092, 2016

37. Slade GD, Sanders AE, Ohrbach R, Fillingim RB, Dubner R, Gracely RH, Bair E, Maixner W, Greenspan JD. Pressure pain thresholds fluctuate with, but do not usefully predict, the clinical course of painful temporomandibular disorder. Pain. 155:2134–2143, 2014

38. Smith SB, Maixner DW, Greenspan JD, Dubner R, Fillingim RB, Ohrbach R, Knott C, Slade GD, Bair E, Gibson DG, Zaykin DV, Weir BS, Maixner W, Diatchenko L. Potential genetic risk factors for chronic TMD: genetic associations from the OPPERA case control study. J Pain. 12:T92–101, 2011

39. Smith SM, Dworkin RH, Turk DC, Baron R, Polydefkis M, Tracey I, Borsook D, Edwards RR, Harris RE, Wager TD, Arendt-Nielsen L, Burke LB, Carr DB, Chappell A, Farrar JT, Freeman R, Gilron I, Goli V, Haeussler J, Jensen T, Katz NP, Kent J, Kopecky EA, Lee DA, Maixner W, Markman JD, McArthur JC, McDermott MP, Parvathenani L, Raja SN, Rappaport BA, Rice ASC, Rowbotham MC, Tobias JK, Wasan AD, Witter J. The Potential Role of Sensory Testing, Skin Biopsy, and Functional Brain Imaging as Biomarkers in Chronic Pain Clinical Trials: IMMPACT Considerations. J Pain. 18:757–777, 2017

40. Stepan L, Shaw CL, Oue S. Temporomandibular disorder in otolaryngology: systematic review. J Laryngol Otol. 131:S50–S56, 2017

41. Sullivan MJL, Bishop SR, Pivik J. The Pain Catastrophizing Scale: Development and validation. Psychological Assessment. 7:524–532, 1995

42. Svensson P, Wang MW, Dong XD, Kumar U, Cairns BE. Human nerve growth factor sensitizes masseter muscle nociceptors in female rats. Pain. 148:473–480, 2010

43. Tan G, Jensen MP, Thornby JI, Shanti BF. Validation of the Brief Pain Inventory for chronic nonmalignant pain. J Pain. 5:133–137, 2004

44. Taylor JM. Psychometric analysis of the Ten-Item Perceived Stress Scale. Psychol Assess. 27:90–101, 2015

45. van de Ruit M, Perenboom MJ, Grey MJ. TMS brain mapping in less than two minutes. Brain Stimul. 8:231–239, 2015

46. van Diepen RM, Mazaheri A. Cross-sensory modulation of alpha oscillatory activity: suppression, idling, and default resource allocation. Eur J Neurosci. 45:1431–1438, 2017

47. Walton KD, Dubois M, Llinas RR. Abnormal thalamocortical activity in patients with Complex Regional Pain Syndrome (CRPS) type I. Pain. 150:41–51, 2010

48. Ware J KM, Dewey J, Gandek B,. How to Score and Interpret Single-Item Health Status Measures: A Manual for Users of the SF-8 Health Survey. QualyMetric. Boston, 2001

